# Hematologic abnormalities after COVID-19 vaccination: A large Korean population-based cohort study

**DOI:** 10.1101/2023.11.15.23298565

**Authors:** Hye Sook Choi, Min-Ho Kim, Myeong Geun Choi, Joo Hun Park, Eun Mi Chun

## Abstract

Adverse hematologic events have been reported after COVID-19 vaccination. The objective of this study was to investigate whether hematologic abnormalities develop after COVID-19 vaccination. Retrospective cohort analyses of data from the Korean National Health Insurance Service (KNHIS) database were conducted from July 2022 to August 2023. We randomly selected data of half of those living in Seoul City as of January 1, 2021 with their diagnostic records up to December 31, 2021. The included participants were vaccinated and nonvaccinated persons aged 20 years or older (n= 4,203,887). Hematologic abnormalities after COVID-19 vaccination were identified as nutritional anemia, hemolytic anemia, aplastic anemia, coagulation defects, and neutropenia using International Classification of Diseases, Tenth Revision codes after index date. Incidence rates of hematologic abnormalities in the vaccination group 3 months after vaccination were significantly higher than those in the nonvaccinated group: 14.79 vs. 9.59 (P<.001) for nutritional anemia, 7.83 vs. 5.00 (P<.001) for aplastic anemia, and 4.85 vs. 1.85 (P<.001) for coagulation defects. COVID-19 mRNA vaccine was associated with higher development of nutritional anemia (odds ratio [OR], 1.230 [95% CI, 1.129-1.339], P<.001) and aplastic anemia (OR, 1.242 [95% CI, 1.110-1.390], P<.001) than the viral vector vaccine. The risk of coagulation defects was increased (OR, 1.986 [95% CI, 1.523-2.589], P<.001) after vaccination, and there was no risk difference between mRNA vaccine and viral vector vaccine (OR, 1.075 [95% CI, 0.936-1.233], P=.306). In conclusions, COVID-19 vaccination increased the risk of hematologic abnormalities. When administering the COVID-19 vaccine, careful observation will be necessary after vaccination.

## 1 INTRODUCTION

The most important weapon against severe acute respiratory syndrome coronavirus 2 (SARS-CoV-2) during the coronavirus 2019 disease (COVID-19) pandemic is vaccination. This is because COVID-19 can be effectively controlled by vaccination of large populations.

To facilitate rapid response, two types of COVID-19 vaccines have been developed earlier; Messenger RNA (mRNA) vaccines including those by Pfizer/BioNTech’s (BNT162b2) and Moderna (mRNA-1273) as well as viral vector vaccines including those by Oxford-AstraZeneca (AZD1222 [ChAdOx1]) and Johnson & Johnson (JNJ-78436735 [Ad26.COV2·S]).^1^ As of August 10, 2023, inactivated, viral vector, mRNA, and protein subunit vaccines have been approved for use by the World Health organization (WHO). ^2^ COVID-19 vaccination is effective in COVID-19 vaccination is effective in preventing severe infection, hospitalization, death, and symptomatic and long COVID-19.^3–5^ While, there are various side effects of COVID-19 vaccinations^6,7^ including immune-mediated disease.^8–12^

The development of vaccine-induced immune thrombosis with thrombocytopenia (VITT) has primarily been considered a serious complication of COVID-19 vaccination.^13^ Most cases of VITT have been associated with vaccines that utilize a recombinant adenoviral vector encoding the spike protein antigen.^14,15^ In addition, VITT following BNT162b2, an mRNA-based vaccine, has been reported.^16^ Autoimmune hemolytic anemia has been reported after mRNA COVID-19 vaccination.^12,17^ Aplastic anemia, which is immune-mediated^18^ and arises after the administration of an mRNA COVID-19 vaccine has been reported.^19–22^ The most commonly reported hematologic abnormalities are secondary immune thrombocytopenia, immune thrombotic thrombocytopenic purpura, autoimmune hemolytic anemia, Evans’ syndrome, and VITT.^23^ Although the incidence of these severe hematologic adverse events is extremely low, their evolution may lead to life-threatening scenarios if treatment is not promptly initiated.

Knowledge of hematologic adverse events related to different COVID-19 vaccines may be beneficial to healthcare providers in assessing and diagnosing the cases earlier. However, there have been no large-scale studies on hematologic abnormalities after COVID-19 vaccination.

We aimed to determine the incidence rates of hematologic abnormalities after COVID-19 vaccination using a reliable real-world large-scale data.

## 2 METHODS

### 2.1 Data source

Korea has implemented a unique national public health insurance system for all its citizens called the Korean National Health Insurance Service (KNHIS). The KNHIS database includes data on all medical treatments and disease codes of the International Classification of Disease, 10th revision (ICD-10). We randomly selected data of half of those living in Seoul City as of January 1, 2021, from the KNHIS database and obtained their 2020-2021 diagnostic records. The diagnosis records included the main and secondary diagnoses, and date hospital visit. Of a total 4,348,412 individuals selected, those 4,203,887 aged >20 years were included.

### 2.2 Study design

Those who received the first vaccination only, died, or had hematologic disease as the primary or secondary diagnosis were excluded from the study. The date of participation in the cohort (index date), was set differently for vaccinated (date of completion of a second vaccination by September 30th, 2021) and non-vaccinated individuals (set as September 30, 2021). Diagnosis records for 365 days prior to the index date were tracked.

### 2.3 Data collection

Viral vector vaccines, mRNA vaccines, and their cross-vaccinations as well as whether or not there was vaccination, were investigated. The covariates included were age, sex, insurance premium level, Charlson Comorbidity Index (CCI), diabetes, hypertension, hyperlipidemia, Chronic obstructive pulmonary disease (COPD), and COVID-19 infection.

KNHIS divides insurance premiums into 20 grades to provide medical benefits as follows: grades 1-6, 7-13, and 14-20 are categorized as low, medium, and high insurance levels, respectively. The diagnosis code for each CCI disease was used, according to Sundararajan et al^24^. Those with each CCI disease (diabetes, hypertension, hyperlipidemia, and COPD) were tracked up using ICD-10 code to 365 days prior to the index date and categorized as patients with 0, 1, or ≥2 observations of primary or secondary disease. COVID-19 was classified as primary or secondary diagnosis prior to the index date, based on ICD-10 code U071.

We investigated nutritional, hemolytic, and aplastic anemia, coagulation defects, and neutropenia as hematologic abnormalities using ICD-10 codes D50–D53, D55–D59, D60-D64, D65-D69, and D70, respectively. The incidence rate of hematologic abnormalities was investigated at 1 week, 2 weeks, 1 month, and 3 months after vaccination.

### 2.4 Study population

Of the 4,203,887 people included in the KNHIS database, 3,839,014 had received COVID-19 vaccine at least once, while 2,154,389 had completed the second vaccination by September 30^th^, 2021, after excluding deaths. The final study sample of vaccination group after excluding those with target disease by the index date was 1,911,617. Of 364,873 nonvaccinated individuals, after excluding deaths by September 30, 2021 and those with target disease by the index date, 324,138 individuals were included (Figure S1).

### 2.5 Statistical analyses

All statistical analyses and data curation were performed using SAS Enterprise Guide (SAS Institute, Cary, NC, USA). Student’s t-test was used to compare continuous variables. The chi-squared test or Fisher’s exact test was used to compare categorical variables. The incidence rate per 10,000 person-years was calculated. Multiple logistic regression model was used to calculate the odds ratios (OR), corresponding 95% confidence intervals (CI), and p-values. Cox regression model was used to calculate the hazard ratio (HR), corresponding 95% CI, and p-value. Statistical significance was set as a two-sided p <0.05. All analyses were performed using only complete data.

## 3 RESULTS

### 3.1 Demographics of the study population

Table 1 presents the demographics of the participants. The vaccination rate was 85.5%. More women (54%) were vaccinated than men (45%) (P<.001). The mean age of the participants was 53 years. The vaccination and non-vaccination groups’ mean ages were 55 and 44 years, respectively (P<.001). Individuals in their 50s, 60s, and 70s had high vaccination rates (91%, 93%, and 94%, respectively) while the rates for those in their 20s,30s, 40s, and 80s, were 76%, 64%, 75%, and 88%, respectively. Vaccination rates were lower in individuals with low (83%) or medium (83%) insurance levels and higher the high insurance level group (87%, P <.001). Those with CCI of 0, 1, or ≥2 had vaccination rates of 81%, 93%, and 93%, respectively. Among patients with diabetes, hyperlipidemia, hypertension, COPD, and COVID-19, respectively, 94%, 95%, 94%, 92%, and 80% were vaccinated.

**TABLE 1.** Participants’ demographics.

|  | Total, No. (%) | Vaccination, No. (%) |  | *p*-value |
| --- | --- | --- | --- | --- |
|  |  | No | Yes |  |
| No./total | 2,235,755 | 324,138 (14.50) | 1,911,617 (85.50) |  |
| Sex |  |  |  |  |
| Male | 1,027,399 (45.95) | 151,921 (46.87) | 875,478 (45.80) | <.001 |
| Female | 1,208,356 (54.05) | 172,217 (53.13) | 1,036,139 (54.20) |  |
| Age, years, mean<br>(SD) | 53.99 (16.97) | 44.69 (16.77) | 55.57 (6.49) | <.001 |
| 20-29 | 263,332 (11.78) | 62,069 (19.15) | 201,263 (10.53) | <.001 |
| 30-39 | 238,700 (10.68) | 84,568 (26.09) | 154,132 (8.06) |  |
| 40-49 | 282,357 (12.63) | 69,200 (21.35) | 213,157 (11.15) |  |
| 50-59 | 530,910 (23.75) | 46,236 (14.26) | 484,674 (25.35) |  |
| 60-69 | 520,452 (23.28) | 32,317 (9.97) | 488,135 (25.54) |  |
| 70-79 | 273,410 (12.23) | 15,018 (4.63) | 258,392 (13.52) |  |
| 80- | 126,594 (5.66) | 14,730 (4.54) | 111,864 (5.85) |  |
| Insurance level |  |  |  |  |
| Low | 569,691 (25.48) | 94,248 (29.08) | 475,443 (24.87) | <.001 |
| Middle | 617,764 (27.63) | 102,021 (31.47) | 515,743 (26.98) |  |
| High | 1,048,300 (46.89) | 127,869 (39.45) | 920,431 (48.15) |  |
| CCI |  |  |  |  |
| 0 | 1,522,302 (68.09) | 278,241 (85.84) | 1,244,061 (65.08) | <.001 |
| 1 | 379,786 (16.99) | 23,756 (7.33) | 356,030 (18.62) |  |
| 2≤ | 333,667 (14.92) | 22,141 (6.83) | 311,526 (16.30) |  |
| Comorbidity |  |  |  |  |
| Diabetes | 323,546 (14.47) | 17,093 (5.27) | 306,453 (16.03) | <.001 |
| Hyperlipidemia | 678,132 (30.33) | 33,541 (10.35) | 644,591 (33.72) | <.001 |
| Hypertension | 605,806 (27.10) | 30,706 (9.47) | 575,100 (30.08) | <.001 |
| COPD | 89,410 (4.00) | 6,267 (1.93) | 83,143 (4.35) | <.001 |
| COVID-19 history | 14,762 (0.66) | 2,916 (0.90) | 11,846 (0.62) | <.001 |

Abbreviations: CCI, Charlson comorbidity index; COPD, chronic obstructive pulmonary disease; COVID-19, Coronavirus disease-2019.
The chi-square test or Fisher's exact test was used for categorical variables, and Student's t-test was used for continuous variables.

### 3.2 Participants’ vaccination history

Those who received mRNA and viral vector vaccines among the first and second vaccination groups were 58% vs. 42% and 64.59% vs. 35.41%, respectively. In the vaccination group, 58.01% and 35.41%, received only mRNA or viral vector vaccines in both the first and second rounds, whereas 6.58% were cross-vaccinated. Vaccination interval occurred over 50.79 days (Table S1).

### 3.3 Rate of development of hematologic abnormalities after vaccination

#### Nutritional anemia

In the vaccinated and nonvaccinated groups, incidence rate of nutritional anemia at 1 week, 2 weeks, 1 month, and 3 months after vaccination were 1.26 vs. 0.71 (P=.006); 2.57 vs. 1.48 (P<.001); 5.12 vs. 3.02 (P<.001); and 14.79 vs. 9.59 (P<.001), respectively (Table 2).

**TABLE 2.**
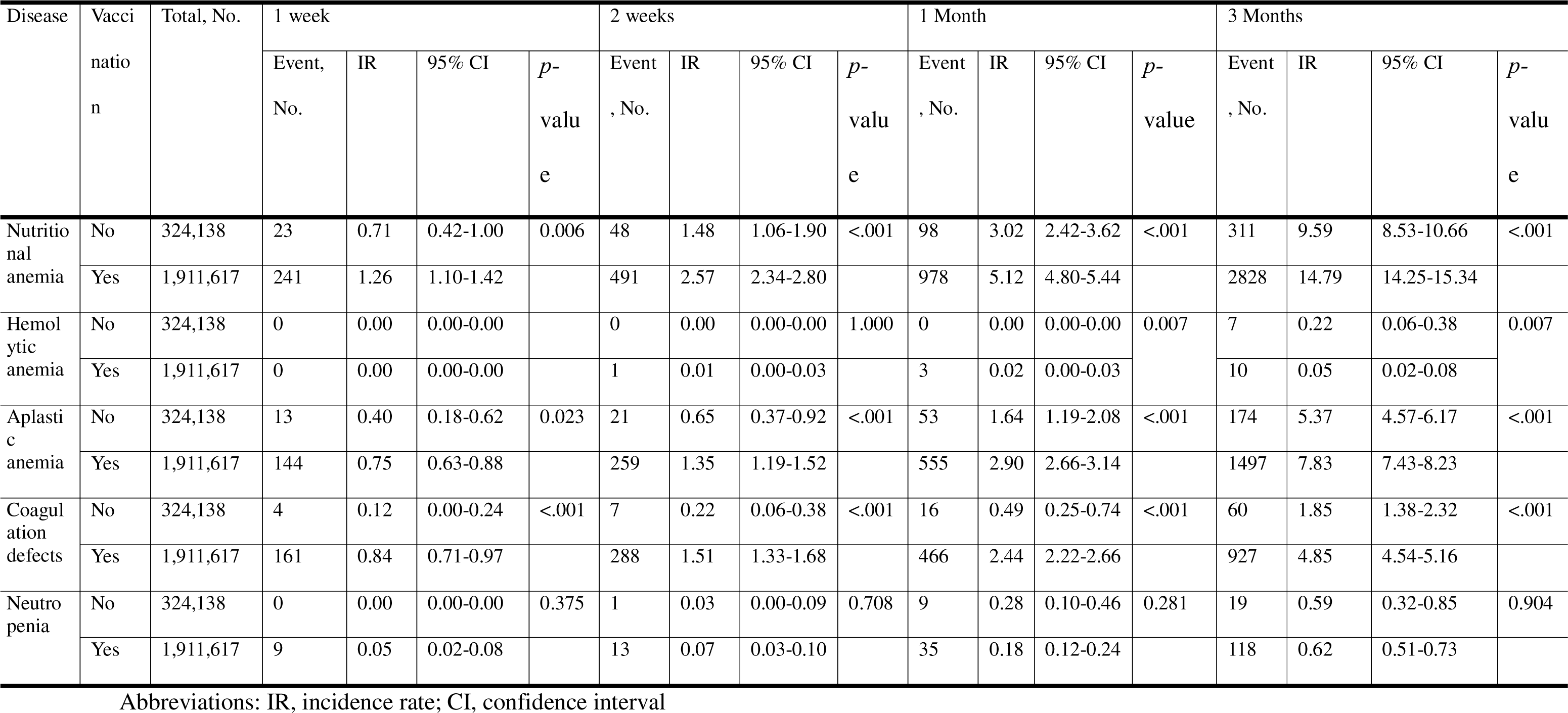
Incidence rate of hematologic abnormalities over time after vaccination.

Multiple logistic regression analyses after adjusting for the covariates (Figure 1) showed that vaccination significantly increased the risk of nutritional anemia at 1 week, 2 weeks, 1 month, and 3 months after vaccination (ORs 1.799 [95% CI, 1.14-2.78, P=.008], 1.713 [95% CI, 126-2.31, P<.001], 1.171 [95% CI, 1.38-2.12, P<.001], and 1.543 [95% CI, 1.36-1.73, P<.001], respectively).

**FIGURE 1.**
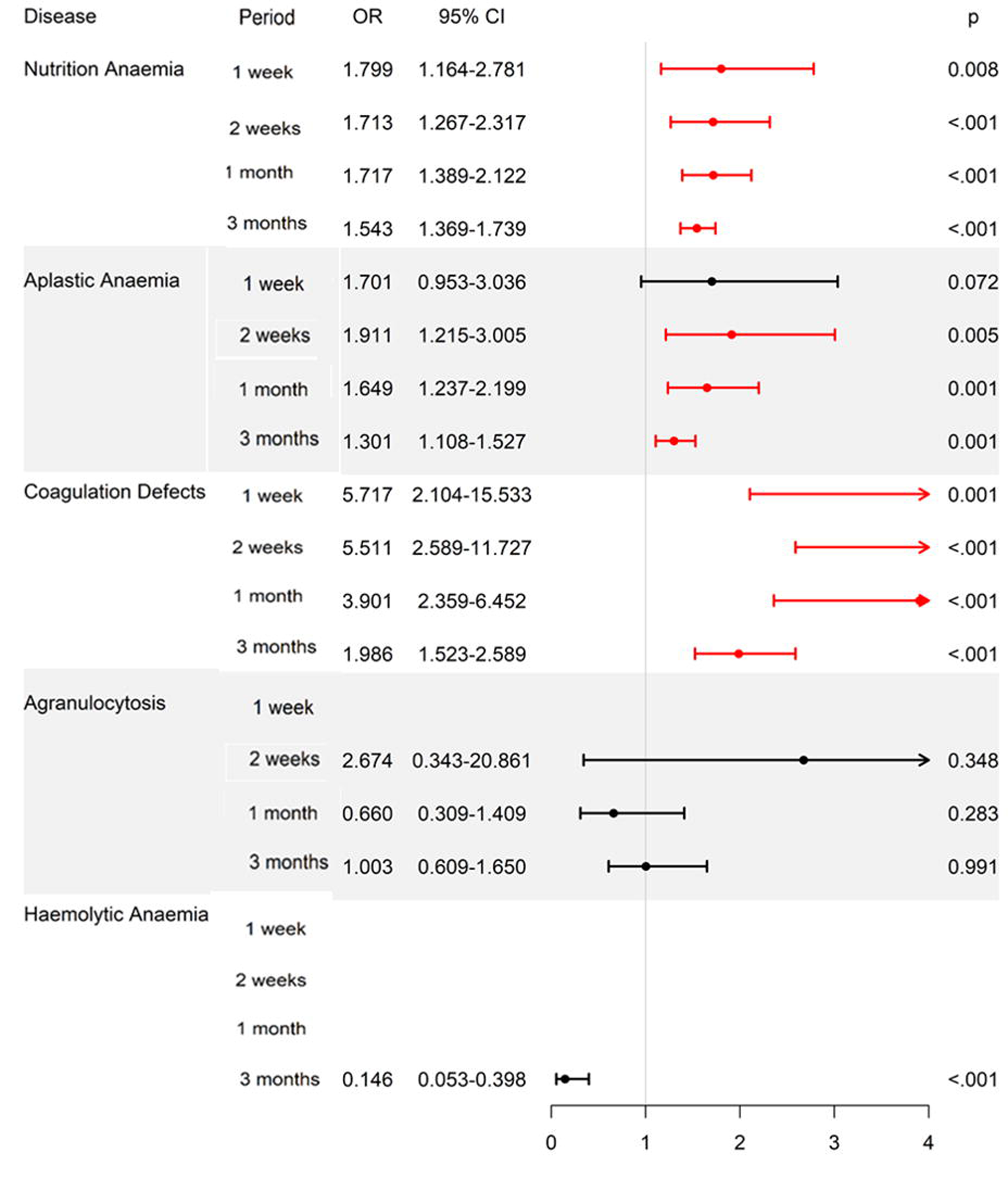
Odd ratio for hematologic abnormalities after vaccination according to the period. Multiple logistic regression analyses showed the risk of development of hematologic abnormalities after vaccination according to the time line. Variables for adjustment: vaccination, gender, age, insurance level, CCI, diabetes, hypertension, hyperlipidemia, COPD, COVID-19 history. CCI, Charlson comorbidity index; COPD, chronic obstructive pulmonary disease; COVID-2019, Coronavirus disease 2019; OR, odd ratio; CI, confidence interval.

Compared to the nonvaccinated group, nutritional anemia 3 months after COVID-19 vaccination showed an HR of 1.601 (95% CI, 1.417-1.809, P<.001) in the mRNA vaccine group; 1.302 (95% CI, 1.136-1.492, P<.001) in the viral vector vaccine group; and 1.775 (95% CI, 1.492-2.110, P<.001) in the cross-vaccination group (Table 3).

**Table 3.** Hazards ratio of hematologic abnormalities at 3 months after COVID-19 vaccination.

| Disease | Variables | Value | HR | 95% CI | *p*-value |
| --- | --- | --- | --- | --- | --- |
| Neutropenia | Vaccine | No |  |  |  |
|  |  | only mRNA vaccine | 0.948 | 0.564-1.592 | 0.839 |
|  |  | only viral vector vaccine | 1.119 | 0.629-1.988 | 0.703 |
|  |  | Cross vaccine | 1.051 | 0.460-2.402 | 0.907 |
|  | Sex | Female | 2.872 | 1.922-4.294 | <.001 |
|  | Age |  | 0.999 | 0.987-1.012 | 0.882 |
|  | Insurance Level | Low |  |  |  |
|  |  | Middle | 1.229 | 0.736-2.052 | 0.430 |
|  |  | High | 1.689 | 1.082-2.637 | 0.021 |
|  | CCI | 0 |  |  |  |
|  |  | 1 | 1.273 | 0.746-2.173 | 0.376 |
|  |  | 2≤ | 2.768 | 1.647-4.652 | <.001 |
|  | Comorbidity | Diabetes | 0.500 | 0.265-0.945 | 0.033 |
|  |  | Hypertension | 0.832 | 0.512-1.353 | 0.459 |
|  |  | Hyperlipidemia | 0.907 | 0.566-1.455 | 0.687 |
|  |  | COPD | 0.774 | 0.329-1.819 | 0.557 |
|  | COVID-19 |  | 2.225 | 0.551-8.981 | 0.261 |

Abbreviations: CCI, Charlson comorbidity index; COPD, chronic obstructive pulmonary disease, COVID-19; Coronavirus disease 2019; HR, hazard ratio of COX regression analysis; CI, confidence interval.

The risk of nutritional anemia increased more significantly after mRNA vaccination than after viral vector vaccination (OR 1.230, 95% CI 1.129-1.339, P<.001). There was no significant difference in the risk of nutritional anemia between the mRNA and cross-vaccination groups. The risk of nutritional anemia increased significantly in the group of cross-vaccination compared with that in the viral vector vaccination only group (OR 1.362, 95% CI 1.168-1.590, P<.001) (Table 4).

**Table 4.** Odd ratio of hematologic abnormalities at 3 months after COVID-19 vaccination according to vaccine types.

| **Disease** | **Reference** | **Values** | **OR** | **95% CI** | **p-value** |
| --- | --- | --- | --- | --- | --- |
| Nutritional anemia | Viral vector vaccine | mRNA vaccine | 1.230 | 1.129-1.339 | <.001 |
|  | mRNA vaccine | Cross vaccine | 1.109 | 0.962-1.277 | 0.155 |
|  | Viral vector vaccine | Cross vaccine | 1.362 | 1.168-1.590 | <.001 |
| Hemolytic anemia | Viral vector vaccine | mRNA vaccine | 3.115 | 0.657-14.760 | 0.152 |
|  | mRNA vaccine | Cross vaccine | Not Available |  |  |
|  | Viral vector vaccine | Cross vaccine | Not Available |  |  |
| Aplastic anemia | Viral vector vaccine | mRNA vaccine | 1.242 | 1.110-1.390 | <.001 |
|  | mRNA vaccine | Cross vaccine | 0.859 | 0.684-1.079 | 0.192 |
|  | Viral vector vaccine | Cross vaccine | 1.067 | 0.840 -1.357 | 0.593 |
| Coagulation defects | Viral vector vaccine | mRNA vaccine | 1.075 | 0.936-1.233 | 0.306 |
|  | mRNA vaccine | Cross vaccine | 1.021 | 0.762-1.372 | 0.886 |
|  | Viral vector vaccine | Cross vaccine | 1.098 | 0.810-1.488 | 0.547 |
| Neutropenia | Viral vector vaccine | mRNA vaccine | 0.847 | 0.567-1.264 | 0.416 |
|  | mRNA vaccine | Cross vaccine | 1.109 | 0.530-2.320 | 0.785 |
|  | Viral vector vaccine | Cross vaccine | 0.939 | 0.431-2.041 | 0.873 |

Abbreviation: OR, odd ratio for logistic regression analysis.

Women had a higher risk of nutritional anemia (HR 2.882, 95% CI 2.650-3.135, P<.001) than men. The risk of nutritional anemia increased with age (HR 1.004, 95% CI, 1.001-1.006, P=.007). The lower the insurance grade, the higher the risk of nutritional anemia in the vaccination group. A CCI score of ≥2 increased the risk of nutritional anemia (HR 1.488, 95% CI 1.314-1.686, P<.001).

#### Hemolytic anemia

The incidence of hemolytic anemia 3 months after COVID-19 vaccination was 7 and 10 in the nonvaccinated and vaccinated groups, respectively (P=.007) (Table 2). The risk of hemolytic anemia at 3 months after COVID-19 vaccination decreased in both mRNA (HR 0.210, 95% CI 0.073-0.602, P=.004) and viral vector vaccination groups (HR 0.068, 95% CI 0.014-0.335, P=.001) significantly compared to the nonvaccinated group. There was no difference in the risk of hemolytic anemia between the mRNA and viral vector vaccine groups (Table 4). Age (HR, 1.047; 95% CI, 1.012-1.083; P=.008) and COVID-19 (HR, 7.741; 95% CI, 1.009-59.400; P=.049) significantly increased hemolytic anemia (Table 3).

#### Aplastic anemia

Incidence rate of aplastic anemia in vaccinated versus nonvaccinated groups were 0.75 vs. 0.40 (P=.023); 1.35 vs. 0.65 (P<.001); 2.90 vs. 1.64 (P<.001); and 7.83 vs. 5.37 (P<.001) at 1 week, 2 weeks, 1 month, and 3 months, respectively, after vaccination (Table 2). The risk of aplastic anemia increased significantly at 2 weeks, 1 month, and 3 months after vaccination (ORs, 1.911 [95% CI, 1.21-3.00, P=.005], 1.649 [95% CI, 1.23-2.19, P=.001], and 1.301 [95% CI, 1.10-1.52, P=.001], respectively) compared to nonvaccinated group (Figure 1).

The risk of aplastic anemia was significantly increased only in the mRNA vaccine group (HR 1.394, 95% CI 1.183-1.643, P<0.001) (Table 3). Compared to the viral vector vaccine group, the mRNA vaccine group had significantly increased risk of aplastic anemia (OR 1.242, 95% CI 1.110-1.390, P<.001) (Table 4).

The risk of aplastic anemia increased with age (HR 1.012, 95% CI 1.007-1.071, P<.001) or in women (HR 1.520, 95% CI 1.334-1.733, P<.001). Female, older age, and CCI ≥2 increased the risk of aplastic anemia significantly (Table 3).

#### Coagulation defects

The incidence of coagulation defects in vaccinated versus nonvaccinated groups were 0.84 vs. 0.12 (P<.001); 0.51 vs. 0.22 (P<.001); 2.44 vs. 0.49 (P<.001); and 4.85 vs. 1.85, P<.001) at 1 week, 2 weeks, 1 month, and 3 months, respectively, after vaccination (Table 2).

The risk of coagulation defects increased significantly at 1 week, 2 weeks, 1 month, and 3 months after vaccination (ORs 5.717 [95% CI, 2.10-15.53, P=.005], 5.511 [95% CI, 2.58-11.72, P=.005], 3.091 [95% CI, 2.35-6.45, P=.005], and 1.986 [95% CI, 1.52-2.58, P=.005], respectively) compared to nonvaccinated group (Figure 1).

The risk of coagulation defects also increased in both the mRNA (HR 2.021, 95% CI 1.542-2.648, P<.001) and viral vector vaccine (HR 1.881, 95% CI 1.419-2.492, P<.001) groups (Table 3). There was no difference in the risk of coagulation defects between the mRNA and viral vector vaccine groups (Table 4).

Female, older age, and CCI 1 or ≥2 increased the risk of coagulation defects significantly. Diabetes decreased the risk of coagulation defects (HR 0.762, 95% CI 0.629-0.924, P=.006) (Table 3).

#### Neutropenia

The neutropenia incidence rate was low (Table 2), and COVID-19 vaccination did not affect the risk of neutropenia (Table 3). The risk of neutropenia increased in women (HR 2.872, 95% CI 1.922-4.294, P<.001) and those with a higher insurance level (HR 1.689, 95% CI 1.082-2.637, P=.021) and CCI≥2 (HR 2.768, 95% CI 1.647-4.652, P<.001) (Table 3).

## 4 DISCUSSION

This study demonstrated the hematologic adverse events associated with COVID-19 vaccination using real-world data. The cumulative incidence rate of nutritional anemia, aplastic anemia, and coagulation defects significantly increased constantly 3 months after the COVID-19 vaccination compared to the nonvaccinated group. We identified that both vaccines were risk factors for nutritional anemia and in particular, mRNA vaccines associated with a higher risk than viral vector vaccines. Additionally, cross-vaccination significantly elevated the risk of nutritional anemia compared to exclusive viral vector vaccination. This suggest that mRNA vaccines may be a particular risk factor for the nutritional anemia, although the exact mechanism is still unknow. Aplastic anemia risk significantly increased only with mRNA vaccine but not with viral vector vaccine. However, another study reported a higher prevalence of hematologic events after viral vector vaccination.^25^ In our study, the risks of nutritional and aplastic anemia were significantly higher in the mRNA vaccine group than in the viral vector vaccine. Similarly, hematologic side effects have been reported with viral vector vaccines^13,14^ as well, but they were reported more frequently after mRNA vaccination.^6,7,9,10,12,15–17,19–21^ Coagulation defects risk increased with both mRNA vaccine and viral vector vaccine, with no difference in risk between the two vaccines. While hemolytic anemia has been reported following COVID-19 vaccination,^17,23,26^ especially with mRNA vaccines,^17,23,26^ our study found that the incidence of hemolytic anemia was low in the COVID-19 vaccine recipient group, regardless of the vaccine type. This suggests the need for further research to confirm the mechanisms between COVID-19 vaccine components and RBC antibodies in a larger population. Only older age and COVID-19 infection increased the risk of hemolytic anemia.

In our data, nutritional anemia risk factors within 3 months after COVID-19 vaccination were vaccine, women, age, lower insurance level, CCI ≥2, and COPD. Aplastic anemia risk factors were vaccine, female sex, increasing age, and CCI ≥2. Coagulation defects risk factors were vaccine, female sex, age, CCI ≥1, hypertension, and hyperlipidemia. Previously, older vaccine recipients had more severe reactions than those aged 18-65,^27^ while younger individuals^28^ were at a higher risk of adverse events. In our study, hyperlipidemia reduced the risk of nutritional anemia, while COPD increased it. Hypertension reduced the risk of aplastic anemia and diabetes reduced the risk of coagulation defects. However, the mechanisms remain unknow; therefore, additional large-scale and laboratory studies are needed. In this study, the vaccine increased the risk of anemia, but interestingly, in a previous real-world cohort study, anemia was associated with lower risk of adverse events after COVID-19 vaccination.^28^

Although several adverse events related to autoimmune diseases have been reported,^29^ there is no established method for assessing the relationship between vaccines and the development of adverse autoimmune diseases. One of the main hypotheses proposed is that vaccine-derived antibodies may have structural similarities to autoantibodies.^30^ This is not unexpected, as the humoral immune response to vaccination involves the generation of antibodies against vaccines that occasionally mimic self-ones.^23^ Platelets, red blood cells, and blood vessels are affected in autoimmune diseases. Thus, it is not surprising that hematologic autoimmune diseases are associated with SARS-CoV-2 vaccines, either mRNA or adenoviral vector-based vaccines. The underlying mechanisms are not completely understood, although the mimicry between viruses and self-antigens plays an important role. It is plausible that if the vaccine-encoded antigen (S protein) in COVID-19 mRNA vaccines enters the human circulation, it can elicit a humoral immune response that contributes to adverse reactions in susceptible individuals.^31^ The hallmark of VITT is the presence of anti-platelet factor 4 autoantibodies, which can trigger platelet activation.^23^ In a reported case of severe aplastic anemia after COVID-19 mRNA vaccination, authors suggested vaccine-derived antibodies may not directly cause it. They proposed that the vaccine antibody titers might provide clues into vaccine-related autoimmune diseases because antibodies against the SARS-CoV-2 spike protein were detected both before and after stem cell transplantation^21^. Multiple factors, including differences in the vaccine itself, genetic susceptibility, and environmental factors are thought to contribute to the development of an immune response. Vaccines, similar to pathogens, can initiate or perpetuate autoimmunity through various mechanisms. The most common mechanism appears to be antigen-induced autoimmunity, including molecular mimicry,^32^ epitope spreading,^33^ bystander activation^34^ and polyclonal activation.^35^ Vaccines mimic the antigens of the targeted pathogen and activate antigen-presenting cells, which stimulate autoreactive T helper cells. These autoreactive T cells and macrophages secrete cytokines, creating a superimposed effect that activates a specific immune response.^30^ Therefore, the immune cells generated by vaccination are similar to those that would be generated by the natural pathogen.^36,37^ Stimulation of T and B cell response through dendritic cell exposure to exogenous mRNA triggers immune response and may play a role in development of multiple sclerosis after vaccination.^38^

The results of this study were based on a large retrospective data; however, the pathophysiologic mechanism underlying the correlation between vaccines and hematologic side effects could not be determined. Nevertheless, the relationship between timing of vaccination and the emergence of hematologic abnormalities raises the possibility of a connection. We suggest the need for careful attention when administering COVID-19 vaccines to people with hematologic diseases. In particular, the older adults, female, and those with underlying diseases require careful observation after vaccination. Prospective studies are needed to establish this connection, comparing RBC-related autoantibodies before and after vaccination, and including unvaccinated control groups.

The limitations of this study include the use of big data based only on the ICD-10 code for hematologic disease as primary or secondary diagnosis. We observed the development of hematologic adverse events by up to 3 months after vaccination, which was a short period. A long-term study to observe the side effects of COVID-19 vaccination longer than one year is needed. As a follow up to this study, we are preparing a study on long term side effects. By September 30, 2021, 2.3 million doses of the mRNA vaccine and 1.4 million doses of the viral vector vaccine had been administered in the study population. This discrepancy makes it difficult to compare the hematologic adverse events among different vaccines.

## 5 CONCLUSIONS

The analyzed data showed that nutritional anemia, aplastic anemia, and coagulation defects increased after COVID-19 vaccination. The risk of nutritional anemia was significantly higher in the mRNA vaccine group than in the viral vector vaccine group. Aplastic anemia risk was significantly increased in mRNA vaccine group but not in viral vector vaccine. The risk of coagulation defects was similarly increased in both mRNA vaccine and viral vector vaccine. Additional risk factors for hematologic abnormalities after COVID-19 vaccination were older age, female sex, and some comorbidities. Further researches are needed to investigate the mechanisms of hematologic abnormalities developing based on the type of COVID-19 vaccines and to conduct long-term outcomes studies.

## AUTHOR CONTRIBUTIONS

Hye Sook Choi designed the study, carried out and interpreted the initial analyses, and drafted and revised the manuscript.

Min-Ho Kim conceptualized and designed the study, carried out and interpreted the initial analyses, and revised the manuscript.

Myeong Geun Choi conceptualized and designed the study, interpreted the initial analyses, and revised the manuscript.

Joo Hun Park conceptualized and designed the study, interpreted the initial analyses, and revised the manuscript.

Eun Mi Chun conceptualized and designed the study, interpreted the initial analyses, and revised the manuscript.

All authors critically revised the manuscript for important intellectual content. All authors approved the final version for submission. The corresponding author attests that all listed authors meet authorship criteria and that no others meeting the criteria have been omitted.

## DATA AVAILABILITY STATEMENT

The data that support the findings of this study are available from the corresponding author upon reasonable request

## SUPPORTING INFORMATION

Additional supporting information can be found online in the Support-ing Information section at the end of this article.

## ETHICAL APPROVAL

The study was approved by the Institutional Review Board of the Ewha Women’s University Hospital (IRB No. EUMC 2022-07-003).

## PATIENT CONSENT

The requirement for informed consent was waived by the IRB of Ewha Women’s University Hospital, and all data were anonymized before retrieval.

## Data Availability

The data that support the findings of this study are available from https://nhiss.nhis.or.kr/bd/ay/bdaya001iv.do upon reasonable request after permission.

https://nhiss.nhis.or.kr/bd/ab/bdaba000eng.do;jsessionid=gz1xQqZdw4em3LDoJmcnMXQrHY0LbeQtqJ8M0F8kB9xaK2hdna2XwOGA1fdl9kE1.primrose22_servlet_engine10

