## Supplementary material for "Hematologic abnormalities after COVID-19 vaccination: A large Korean population-based cohort study": FIGURE S1

**Supplemental FIGURE 1**

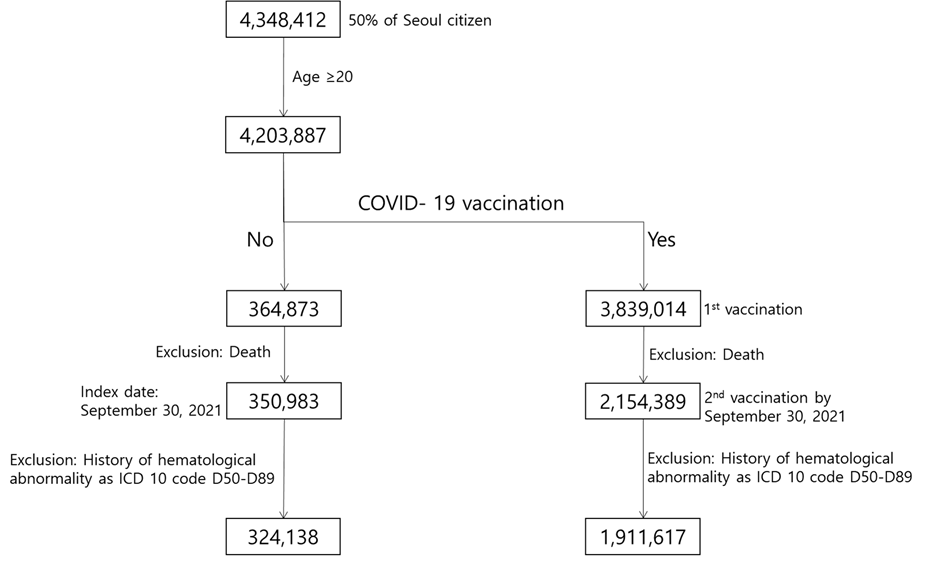

**FIGURE S1.** Flow chart of the study population. Population is 50% of Seoul citizens from KNIHS database.

Abbreviation: ICD, international classification of disease.
