## Supplementary material for "Hematologic abnormalities after COVID-19 vaccination: A large Korean population-based cohort study": TABLE S1

**TABLE S1.** History of vaccination of the subjects.

|  | Total, No. (%) | Vaccination, No. (%) | | *P* value |
| --- | --- | --- | --- | --- |
|  |  | No | Yes |  |
| Total | 2,235,755 (100) | 324,138 (14.50) | 1,911,617 (85.50) |  |
| 1^st^ vaccine |  |  |  |  |
| mRNA vaccine | 1,108,869 (58) |  | 1,108,869 (58) |  |
| BNT162b2 | 1,080,301 (56.51) |  | 1,080,301 (56.51) |  |
| mRNA-1273 | 28,568 (1.49) |  | 28,568 (1.49) |  |
| Viral vector vaccine | 80.748 (42) |  | 80.748 (42) |  |
| ChAdOx1 | 802,736 (41.99) |  | 802,736 (41.99) |  |
| Ad26.COV2·S | 12 (0.00) |  | 12 (0.00) |  |
| 2^nd^ vaccine |  |  |  |  |
| mRNA vaccine | 1,234,696 (64.59) |  | 1,234,696 (64.59) |  |
| BNT162b2 | 1,206,108 (63.09) |  | 1,206,108 (63.09) |  |
| mRNA-1273 | 28,588 (1.50) |  | 28,588 (1.50) |  |
| Viral vector vaccine | 676,921 (35.41) |  | 676,921 (35.41) |  |
| ChAdOx1 | 676,915 (35.41) |  | 676,915 (35.41) |  |
| Ad26.COV2·S | 6 (0.00) |  | 6 (0.00) |  |
| Cross-vaccine |  |  |  |  |
| BNT162b2-BNT162b2 | 1,080,276 (56.51) |  | 1,080,276 (56.51) |  |
| BNT162b2-mRNA-1273 | 22 (0.00) |  | 22 (0.00) |  |
| BNT162b2-ChAdOx1 | 2 (0.00) |  | 2 (0.00) |  |
| BNT162b2-Ad26.COV2·S | 1 (0.00) |  | 1 (0.00) |  |
| mRNA-1273-BNT162b2 | 3 (0.00) |  | 3 (0.00) |  |
| mRNA-1273-mRNA-1273 | 28,565 (1.49) |  | 28,565 (1.49) |  |
| ChAdOx1-ChAdOx1 | 676,907 (35.41) |  | 676,907 (35.41) |  |
| ChAdOx1- BNT162b2 | 125,823 (6.58) |  | 125,823 (6.58) |  |
| ChAdOx1- mRNA-1273 | 1 (0.00) |  | 1 (0.00) |  |
| ChAdOx1-Ad26.COV2·S | 5 (0.00) |  | 5 (0.00) |  |
| Ad26.COV2·S-ChAdOx1 | 6 (0.00) |  | 6 (0.00) |  |
| Ad26.COV2·S-BNT162b2 | 6 (0.00) |  | 6 (0.00) |  |
| Cross-vaccination |  |  |  |  |
| No | 324,138 (14.50) | 324,138 (100.00) |  |  |
| only mRNA vaccine | 1,108,873 (49.60) |  | 1,108,873 (58.01) |  |
| only viral vector vaccine | 676,907 (30.28) |  | 676,907 (35.41) |  |
| cross | 125,837 (5.63) |  | 125,837 (6.58) |  |
| Vaccination interval, days, mean (SD) | 50.79 (23.36) |  | 50.79 (23.36) |  |

SD, standard deviation.
